# A Linear Mixed Model with Measurement Error Correction (LMM-MEC): A Method for Summary-data-based Multivariable Mendelian Randomization

**DOI:** 10.1101/2023.04.25.23289099

**Authors:** Ming Ding, Fei Zou

## Abstract

Summary-data-based multivariable Mendelian randomization (MVMR) methods, such as MVMR-Egger, MVMR-IVW, MVMR median-based, and MVMR-PRESSO, assess the causal effects of multiple risk factors on disease. However, accounting for variances in summary statistics related to risk factors remains a challenge. We propose a linear mixed model with measurement error correction (LMM-MEC) that accounts for the variance of summary statistics for both disease outcomes and risk factors. In step I, a linear mixed model is applied to account for the variance in disease summary statistics. Specifically, if heterogeneity is present in disease summary statistics, we treat it as a random effect and adopt an iteratively re-weighted least squares algorithm to estimate causal effects. In step II, we treat the variance in the summary statistics of risk factors as multiple measurement errors and apply a regression calibration method for simultaneous multiple measurement error correction. In a simulation study, when using independent genetic variants as instrumental variables (IV), our method showed comparable performance to existing MVMR methods under conditions of no pleiotropy or balanced pleiotropy with the outcome, and it exhibited higher coverage rates and power under directional pleiotropy. Similar findings were observed when using genetic variants with low to moderate linkage disequilibrium (LD) (0 < *ρ*^2^ ≤ 0.3) as IVs, although coverage rates reduced for all methods compared to using independent genetic variants as IVs. In the application study, we examined causal associations between correlated cholesterol biomarkers and longevity. By including 739 genetic variants selected based on P values <5×10^-5^ from GWAS and allowing for low LD (*ρ*^2^ ≤ 0.1), our method identified that large LDL-c were causally associated with lower likelihood of achieving longevity.

## 1. INTRODUCTION

In recent years, genome-wide association studies (GWAS) with numerous phenotypes have been conducted, and the summary statistics generated provide us with an invaluable opportunity to assess causality between phenotypes and the disease using Mendelian randomization (MR). The inferred causal associations of MR methods can be affected by four factors: 1) the strength of instrumental variables (IVs), indicated by the F statistic between a genetic variant and a risk factor; 2) pleiotropy, where one variant may have direct effects on multiple risk factors or the disease outcome; 3) the independence of IVs, assessed by linkage disequilibrium (LD) between genetic variants; and 4) variance in summary statistics in relation to both risk factors and the disease.

Univariate MR approaches that examine the causal effect of a single risk factor typically recognize pleiotropy as a limitation or attempt to detect its presence based on distributional properties. For example, MR-IVW calculates an average causal estimate across all genetic variants using inverse-variant weighted meta-analysis, while acknowledging that pleiotropy may manifest as heterogeneity.^1^ Other methods are designed to detect and exclude instruments inferred to have potential pleiotropic influences (e.g. MR-median,^1^ MR-PRESSO,^2^ GSMR with HEIDI^3,4^). Notably, by including an intercept term, MR-Egger is robust to pleiotropy with the disease under the InSIDE (INstrument Strength Independent of Direct Effect) assumption.^5^ Still other univariate MR methods, distinguish possible direct and indirect effects (e.g. MRMix^6^), estimate the proportion of instrumental effects that are causal as opposed to pleiotropic (e.g. LCV^7^), or using Bayesian approaches to separate causal from confounding effects (e.g. CAUSE^8^). However, all of these methods were developed under the NOME (NO Measurement Error) assumption,^9^ where the variance of summary statistics with the risk factor is ignored. To deal with this, methods have been developed to detect outliers in the presence of weak instruments (e.g. radial MR using modified second-order weights^9^), or to account for random error in summary statistics related to the risk factor (e.g. MR-RAPS,^10^ MR-MtRobin^11^).

Given the many methods developed, an intrinsic limitation of univariate MR methods is that they do not explicitly model pleiotropy with other risk factors. To address this, multivariable MR (MVMR) methods have been proposed, which regress summary statistics of disease on those of multiple risk factors, with coefficients indicating causal associations between risk factors and disease.^12-15^ The MR-Egger,^16^ MR-IVW,^17^ median-based MR,^17^ and MR-PRESSO methods have been extended to the multivariable case. However, few MVMR methods have accounting for the variances of summary statistics with risk factors. In this paper, we propose a linear mixed model with measurement error correction (LMM-MEC) that accounts for the variance of summary statistics not only in the disease outcome but also in risk factors. In step I, a linear mixed model is applied, treating the variance in disease summary statistics as fixed or random effects. Specifically, if heterogeneity is present in disease summary statistics, we treat it as a random effect and adopt an iteratively re-weighted least squares algorithm to estimate causal effects.^18^ In step II, we treat variance in summary statistics of risk factors as multiple measurement errors and apply a regression calibration method for simultaneous measurement error correction.^19-22^

We evaluated the robustness of our model to balanced and directional pleiotropy with the outcome under the InSIDE assumption. Here, balanced pleiotropy is defined as the average direct pleiotropy across genetic variants equaling zero, while directional pleiotropy is defined as the average direct pleiotropy across genetic variants differing from zero. We assessed the robustness of our model to the strong IV assumption. As our method accounts for correlations between genetic variants in both steps, we also evaluated the robustness of our model to correlated genetic variants by allowing low to moderate LD.

## 2. METHOD DEVELOPMENT

### 2.1. Overview of the Summary-data-based MVMR

As shown in **Figure 1a**, we use *N* genetic variants, *G*_*n*_ (*n* = 1, …, *N*) as IVs to examine the causal associations between *M* risk factors *X*_*m*_ (*m* = 1, …, *M*) and disease Y, with unknown confounding U. We allow the IVs to be associated with one or more *X*_*m*_*s* or directly associated with Y. With participant-level data for *G*_*n*_*s* and *X*_*m*_s, we link them with each other and U and Y via the following models:

**Figure 1.**
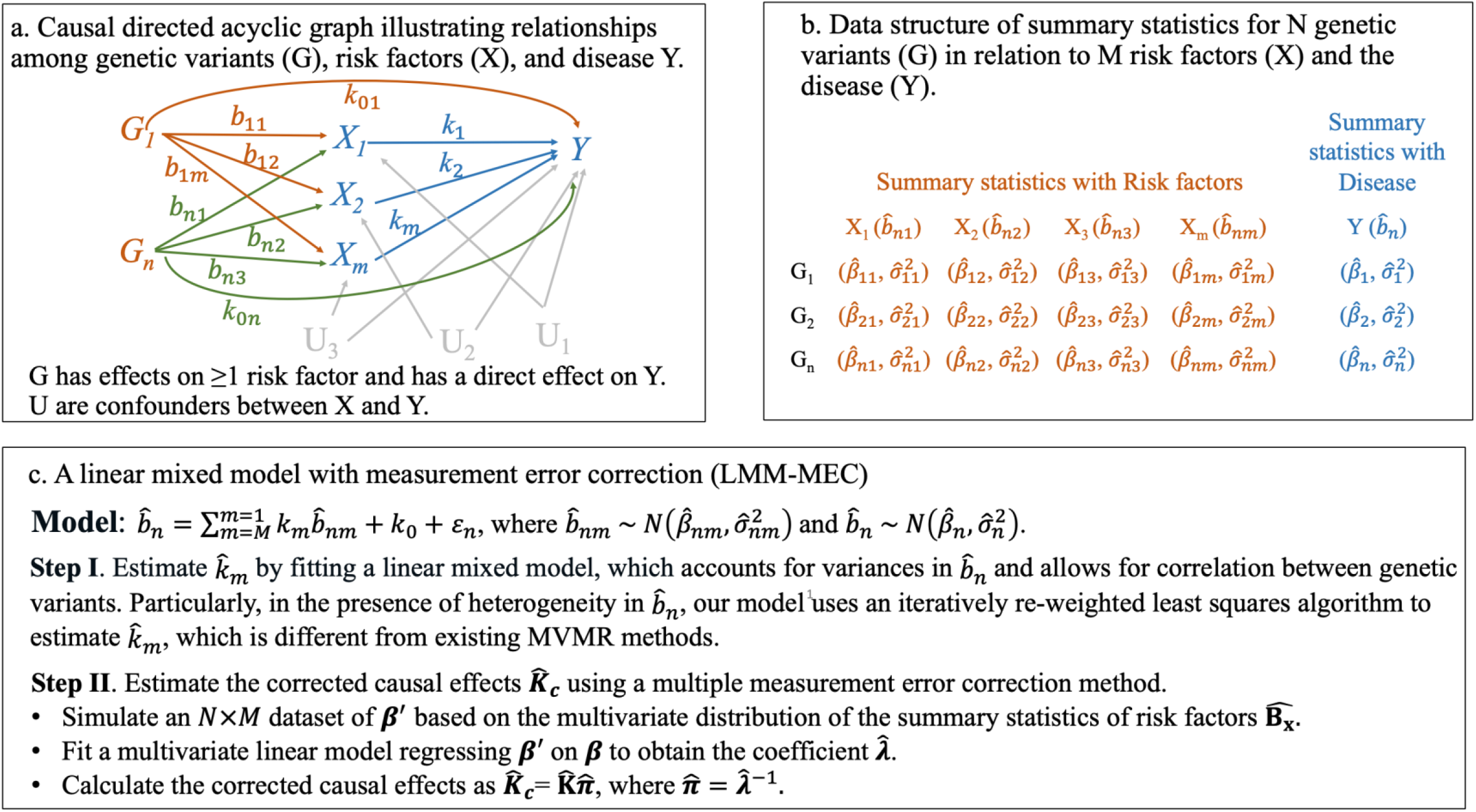
An illustration of the workflow of the linear mixed model with measurement error correction (LMM-MEC).

**Model 1**. 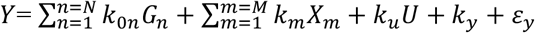

**Model 1**. 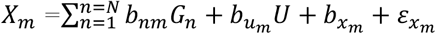

where *b*_*nm*_ is the effect of *G*_*n*_ on *X*_*m*_; *k*_*u*_ and 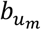 are the effects of confounder U on Y and *X*_*m*_, respectively. *k*_*y*_ and 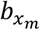 are intercepts, and *ε*_*y*_ and 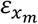 are random errors with normal distributions. *k*_0*n*_ is the direct effect of *G*_*n*_ on *Y. k*_*m*_ is the causal effect of risk factor *X*_*m*_ on *Y*, the parameter that we wish to estimate.

It then follows that 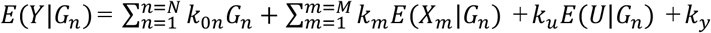, where 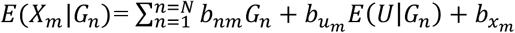. As G and U are assumed to be independent, *E*(*U*|*G*_*n*_) =0, thus,

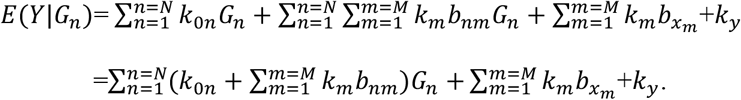

With participant-level data, we can regress Y on *G*_*n*_s with the following model:

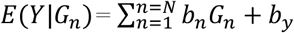

where 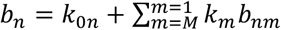. To estimate the causal effect *k*_*m*_, we can utilize existing summary statistics of genetic variants in relation to risk factors 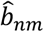 and summary statistics with the outcome 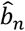 (**Figure 1b**). Specially, 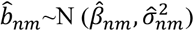, where 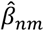 is the estimated effect size of genetic variant *n* on risk factor *m* and 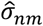 is the standard error from GWAS with risk factor *m*. 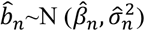, where 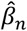 is the effect size of genetic variant *n* on the disease outcome and 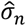 is the standard error from GWAS with the disease outcome.

When *G*_*n*_*s* are not in LD, the following linear regression model can be fitted:

**Model 3**. 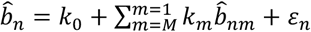, where the intercept *k*_0_ represents the average direct effect of *k*_0*n*_ across all genetic variants.

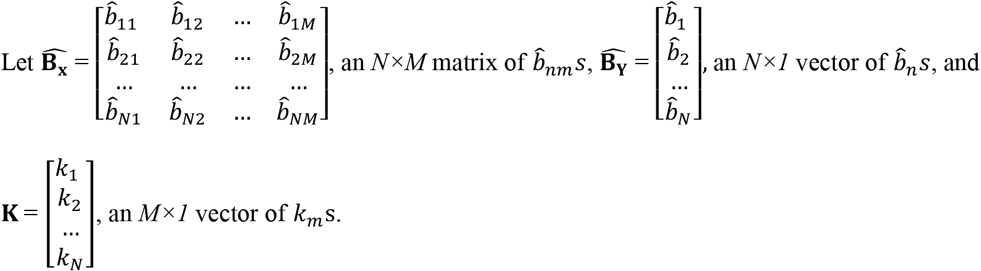

**Model 3** can then be expressed in matrix form as 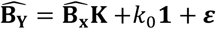.

Assuming no LD among *G*_*n*_*s*, summary statistics 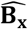 and 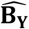 are consistent estimates of **B**_**x**_ and **B**_**Y**_, respectively, where **B**_**x**_ are the true associations of genetic variants with risk factors and **B**_**Y**_ are the true associations of genetic variants with the disease outcome. Under the NOME assumption that there is no measurement error in 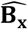 (i.e., the standard error of 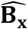 is assumed to be 0),^9^ the 2-Stage Least Squares (2SLS) weighted regression estimator of **K** is given as

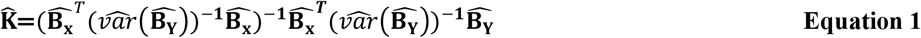

with an associated covariance estimator

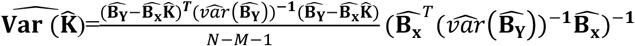, where 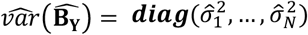 and 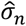 is the standard error of 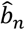.

When *G*_*n*_*s* are in LD, following Sanderson *et al* (2021),^23^ **Equation 1** still applies with the diagonal matrix 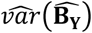 being replaced by

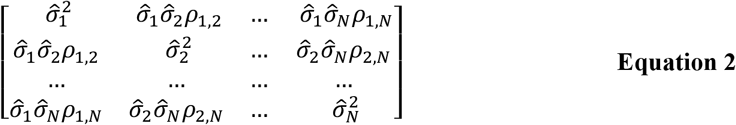

where *ρ*_*r*,*s*_ is the correlation between summary statistics 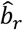 and 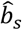. According to Burgess et al (2016),^24^ *ρ*_*r*,*s*_ can be approximated by the square root of the LD between the two genetic variants *r* and *s*.

### 2.2. Our Approach: LMM-MEC

Compared to existing MVMR methods, our approach offers two key improvements. First, our method accounts for heterogeneity effects of genetic variants on the disease outcome while allowing for LD among them through a linear-mixed model with an iteratively re-weighted least squares algorithm to obtain 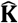 an estimate of **K**.^18^ Second, in deriving 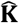 it is often assumed that *b*_*nm*_ is estimated accurately, i.e., 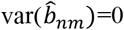 which never holds in practice. We treat 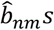 as variables with measurement errors which we simultaneously account for using multiple measurement error correction methods.^19^ An illustration of the workflow of the LMM-MEC work is shown in **Figure 1c**. In step I, we estimate the causal effects 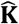 allowing for heterogeneity effects of genetic variants on the disease outcome. In step II, we obtain corrected causal effects using multiple measurement error correction methods.

#### 2.2.1. Step I of LMM-MEC

We assess heterogeneity effects of genetics variants to disease outcome by calculating Cochran’s Q statistic as follows:

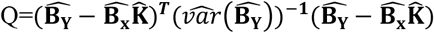 and 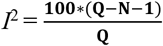, with *I*^2^ > 50% indicating significant heterogeneity.^25^ If no heterogeneity is found, 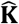 can be estimated using the 2SLS estimator. In the presence of heterogeneity, we add 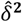 to 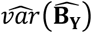 in **Equation 2**, that is

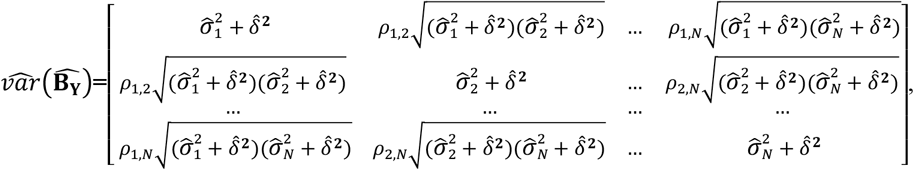

We then use an iteratively re-weighted least squares algorithm to estimate 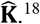 Initially, we assume 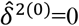 and use the 2SLS estimator to obtain 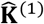. In the next loop, 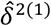 can be estimated as:

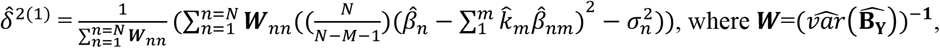

***W***_*nn*_ is the *n*^th^ row and *n*^th^ column of the matrix ***W***, 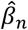 is the mean value of the summary statistics 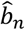, and 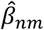 is the mean value of the summary statistics 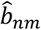.

The absolute value of 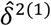 is used if the estimate is negative. We iteratively estimate 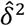 and 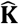 until the estimate of 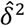 converges.

#### 2.2.2. Step II of LMM-MEC

In Step I, 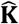 is obtained under the NOME assumption. However, 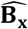 are summary statistics, and ignoring their estimation variances can lead to inaccurate estimates of 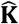 In this step, we treat 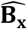 as covariates with random measurement errors, and apply a regression calibration method for adjusting their measurement errors, which is a well-established approach in epidemiology.^19-22^ To obtain the corrected causal effect estimate of **K**, denoted as 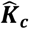, we follow Rosner *et al* (1990)^20^, where 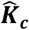 is adjusted by a correction factor 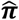 as 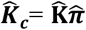.To get 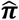 we employ the following simulation strategy.

First, we simulate a validation data by randomly generating an *N* × *M* matrix 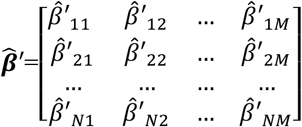 from a multivariate normal distribution 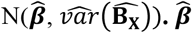 is the mean value of 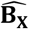 and can be written as 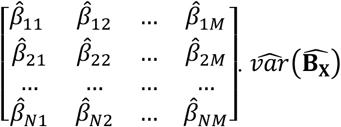 is a matrix with (*N×M*) rows and (*N×M*) columns and can be written as

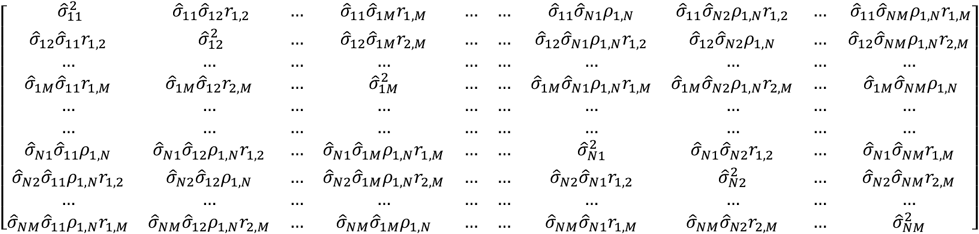

*ρ*_*rs*_ is the Pearson correlation between genetic variants *r* and *s*, and *r*_*ij*_ is the correlation between summary statistics of risk factors *i* and *j*, which can be estimated using phenotype correlation from GWAS studies or from a dataset with a similar structure.^23^

Second, apply a regression calibration model. In the validation study, a multivariate linear model is fit by regressing 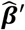 on 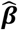 where the coefficient 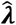 is an *M* × *M* matrix estimated as 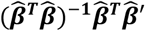 and 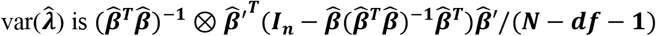, with *⊗* denoting Kronecker product, ***I***_***n***_ as the identity matrix, and *df* as the degrees of freedom of the regression model. We simulate the validation study multiple times and obtain 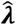 each time. The 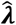 estimates are then pooled using Rubin’s rule.^26^

Finally, 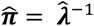. The corrected causal effect 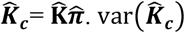 is an *M* × *M* matrix, and the *p*^th^ row and *q*^th^ column of the matrix of var 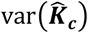 can be calculated as follows.^27^

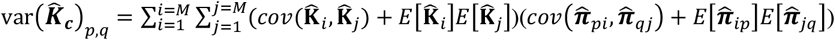 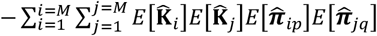, where 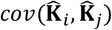 is the covariance between 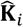 and 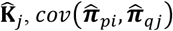 is the covariance between 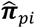 and 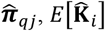 is the mean value of 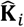 and 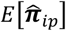 is the mean value of 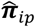.

## 3. SIMULATION STUDY

### 3.1. Simulation of Datasets

We performed a simulation study to validate the utility of our model. For each dataset, we generated data on 100 genetic variants (G) from 2000 participants. Alleles of genetic variants were simulated from standard multivariate normal distributions, with covariance representing the Pearson correlation between genetic variants. Each allele was simulated twice and recoded into 0 and 1 using a cutoff from a uniform distribution *U* [0, 0.8] to generate genetic variants with a minor allele frequency ranging from 0.2 to 0.5. The two alleles were summed to generate a genetic variant coded as 0, 1, and 2. We generated risk factor *m* (*X*_*m*_) and outcome *Y* from **Models 4-5**.

**Model 4:**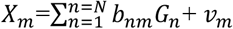, where *v* _*m*_ ∼*N* (0,1) and *b*_*nm*_ was simulated from multivariate normal distributions with covariance indicating correlation between genetic variants.

**Model 5:**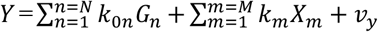, where *v* _*y*_ ∼*N* (0,1), *k*_0*n*_ represents the direct effect of genetic variant *n* on Y, and *k*_*m*_ denotes the causal effect of risk factor *X*_*m*_ on Y.

To test the validity of our model in accurately estimating causal effects, we simulated three risk factors, *X*_1_, *X*_2_, *X*_3_, which had positive (*k*_1_), null (*k*_2_), and negative (*k*_3_) effects on Y. To evaluate the robustness of our model to LD between genetic variants, we simulated scenarios with no (*ρ*^2^=0), low (0<*ρ*^2^<0.1), and moderate (0.1<*ρ*^2^<0.3) LD by changing the covariance of the multivariate normal distributions from which the alleles were sampled. To assess the robustness of our model against pleiotropy with the outcome, we simulated scenarios with no pleiotropy, balanced pleiotropy, and directional pleiotropy while satisfying the InSIDE assumption. To examine the robustness of our model to weak IVs, we included a variable ξ in generating 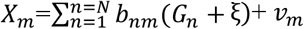, where ξ was simulated from a normal distribution with mean 0 and variance varying from 0.1 to 10. To show that our model can be applied to the univariate MR case, we simulated only one risk factor (X) each time, with positive, negative, and null effects on Y.

To fit MVMR models, we randomly split the 2000 participants into two subsets, each containing 1000 participants. One subset was used to obtain summary statistics with the risk factors, and the other subset was used for the outcome. We employed our model to estimate the causal effects and compared our results to those from MRMV-IVW, MRMV-Egger, MRMV-median method, and MVMR-PRESSO.^2,28^ In the application of our model, in step I, we assessed heterogeneity in the summary statistics related to disease across genetic variants by calculating Cochran’s Q statistic and *I*^2^, with *I*^2^ < 50% indicating no heterogeneity.^25^ In step II, we simulated the validation study for measurement error correction for 100 times. For each risk factor, we calculated the mean F statistic across genetic variants and the conditional F statistic accounting for correlation with other risk factors.^23^ In the univariate setting, we further compared our model to radial IVW and radial Egger methods, which use modified second-order weights to estimate causal effects.^9^ We conducted simulations 1000 times, calculating the average of the estimated causal effects along with their standard errors, coverage, and power across the 1000 datasets. Coverage was defined as the proportion of 95% CIs including the true causal effect across all simulated datasets. Power was estimated as the proportion of 95% CIs that excluded zero in settings with a significant causal effect. We further evaluated the performance of our model by increasing the sample size to 10,000 and 20,000.

### 3.2. Simulation Results

We tested the robustness of our model to the strong IV using independent genetic variants as IVs. The coverage rate and power remained high until the mean F statistics was lowered to 10, after which they gradually decreased when the mean F statistics fell below 10 (**Figure 2**). There was a drastic decrease in coverage and power when the mean F statistic dropped below 5. These trends were observed under the conditions of no pleiotropy, balanced pleiotropy, and directional pleiotropy. Thus, in **Table 1**, we simulated strong IVs to assess model performance, yielding a mean F statistic of 17.3 and a conditional mean statistic of 16.8. Our method is robust to pleiotropy: It showed performance comparable to existing MVMR methods under the conditions of no pleiotropy with the outcome and balanced pleiotropy, and exhibited higher coverage rates under directional pleiotropy, with coverage rates 90%, 96%, and 91% for *k*_1_, *k*_2_, *k*_3_, respectively (**Table 1**). We applied our model by increasing the sample size from 5000 to 20000, which resulted in improved coverage rates to 95% for *k*_1_, *k*_2_, and *k*_3_ under conditions of no pleiotropy, balanced pleiotropy, and unbalanced pleiotropy (**Table S1**).

**Table 1.**
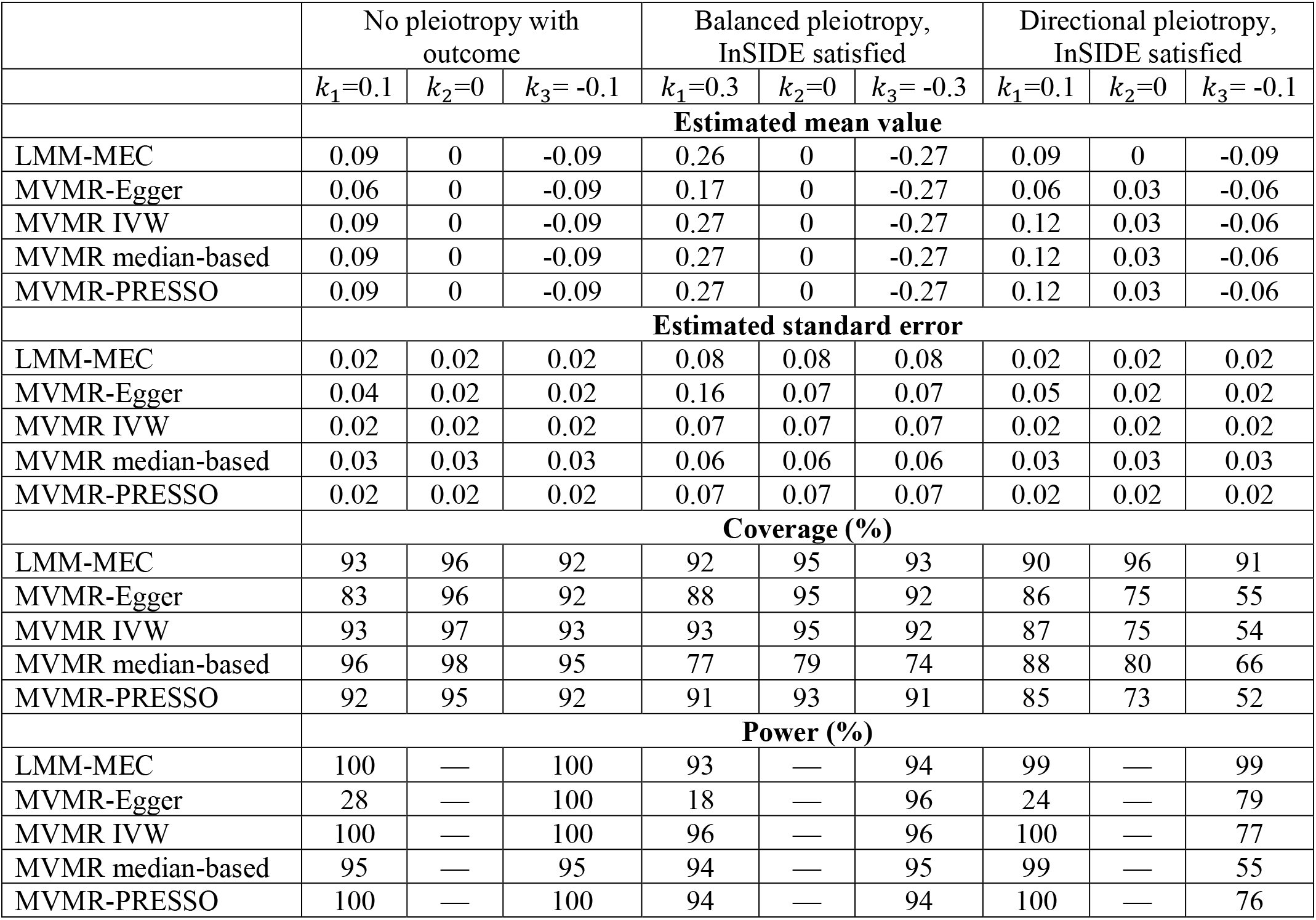
A simulation study estimating causal associations between positive (X_1_), null (X_2_), and negative (X_3_) risk factors and outcome (Y) under no linkage disequilibrium (*ρ*^2^=0) between genetic variants.

**Figure 2.**
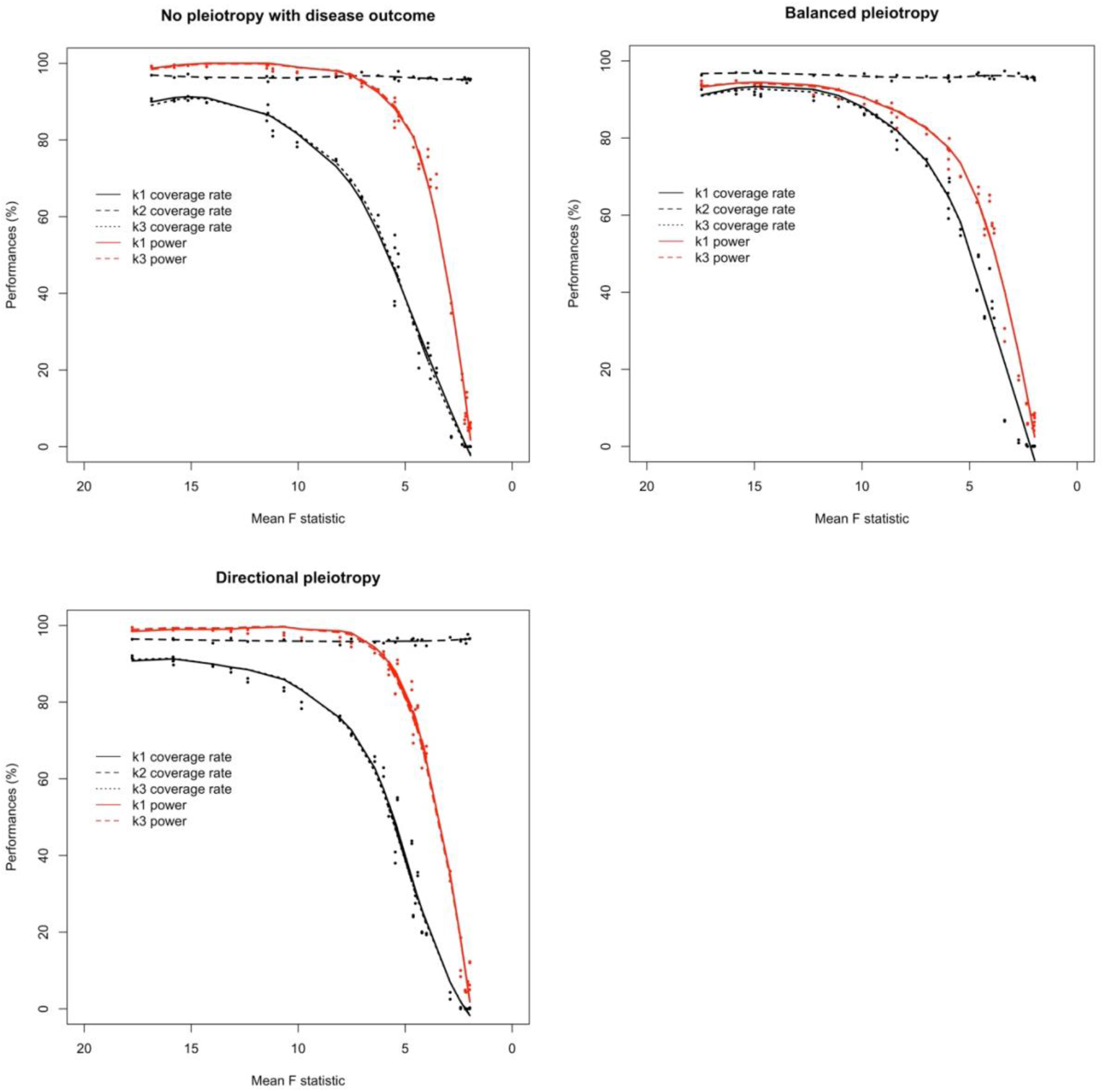
Simulation studies estimating causal associations between positive (X_1_), null (X_2_), and negative (X_3_) risk factors and outcome (Y) using LMM-MEC method according to strengths of instrument variables under no linkage disequilibrium (*ρ*^2^ = 0) between genetic variants.

We simulated genetic variants with low to moderate LD (0 < *ρ*^2^ ≤ 0.3) as IVs, and compared our model to MVMR-Egger and MVMR-IVW which account for LD (using the “mr_mvegger” and “mr_mvivw” packages). Our method showed comparable performance under the condition of no pleiotropy, and higher coverage rates than MVMR-Egger and MVMR-IVW under balanced and directional pleiotropy (**Table 2**). However, the coverage rates reduced for all methods when using correlated IVs compared to independent IVs. We further evaluated the coverage rates of our model by increasing the sample size from 5000 to 20000. The coverage rates improved to 95% under no pleiotropy but were below 95% under the conditions of balanced and directional pleiotropy (**Table S1**).

**Table 2.** A simulation study estimating causal associations between positive (X_1_), null (X_2_), and negative (X_3_) risk factors and outcome (Y) under low to moderate linkage disequilibrium (0 < *ρ*^2^ ≤ 0.3) between genetic variants.

|  | No pleiotropy with outcome |  |  | Balanced pleiotropy, InSIDE satisfied |  |  | Directional pleiotropy, InSIDE satisfied |  |  |
| --- | --- | --- | --- | --- | --- | --- | --- | --- | --- |
| | $k_1=0.1$ | $k_2=0$ | $k_3=-0.1$ | $k_1=0.3$ | $k_2=0$ | $k_3=-0.3$ | $k_1=0.1$ | $k_2=0$ | $k_3=-0.1$ |
| Low LD ($0 < \rho^2 \leq 0.1$) | | | | | | | | | |
|  | **Estimated mean value** |  |  |  |  |  |  |  |  |
| LMM-MEC | 0.09 | 0 | -0.09 | 0.26 | 0 | -0.26 | 0.1 | 0.01 | -0.08 |
| MVMR-Egger | 0.09 | 0 | -0.09 | 0.26 | 0 | -0.26 | 0.11 | 0.03 | -0.06 |
| MVMR IVW | 0.09 | 0 | -0.09 | 0.26 | 0 | -0.26 | 0.18 | 0.09 | 0 |
|  | **Estimated standard error** |  |  |  |  |  |  |  |  |
| LMM-MEC | 0.02 | 0.02 | 0.02 | 0.08 | 0.08 | 0.08 | 0.02 | 0.02 | 0.02 |
| MVMR-Egger | 0.02 | 0.02 | 0.02 | 0.08 | 0.08 | 0.08 | 0.03 | 0.03 | 0.03 |
| MVMR IVW | 0.02 | 0.02 | 0.02 | 0.08 | 0.08 | 0.08 | 0.03 | 0.03 | 0.03 |
|  | **Coverage (%)** |  |  |  |  |  |  |  |  |
| LMM-MEC | 90 | 97 | 92 | 90 | 92 | 86 | 96 | 95 | 84 |
| MVMR-Egger | 91 | 97 | 93 | 89 | 83 | 80 | 93 | 73 | 65 |
| MVMR IVW | 90 | 97 | 92 | 58 | 61 | 63 | 29 | 21 | 13 |
|  | **Power (%)** |  |  |  |  |  |  |  |  |
| LMM-MEC | 99 | — | 99 | 90 | — | 88 | 97 | — | 89 |
| MVMR-Egger | 99 | — | 99 | 88 | — | 86 | 97 | — | 69 |
| MVMR IVW | 100 | — | 100 | 76 | — | 79 | 100 | — | 16 |
| Moderate LD ($0.1 < \rho^2 \leq 0.3$) | | | | | | | | | |
|  | **Estimated mean value** |  |  |  |  |  |  |  |  |
| LMM-MEC | 0.09 | 0 | -0.08 | 0.25 | 0 | -0.26 | 0.11 | 0.02 | -0.06 |
| MVMR-Egger | 0.09 | 0 | -0.08 | 0.25 | 0 | -0.26 | 0.11 | 0.02 | -0.06 |
| MVMR IVW | 0.09 | 0 | -0.09 | 0.26 | -0.01 | -0.27 | 0.2 | 0.11 | 0.02 |
|  | **Estimated standard error** |  |  |  |  |  |  |  |  |
| LMM-MEC | 0.02 | 0.02 | 0.02 | 0.09 | 0.09 | 0.09 | 0.03 | 0.03 | 0.03 |
| MVMR-Egger | 0.02 | 0.02 | 0.02 | 0.09 | 0.09 | 0.09 | 0.03 | 0.03 | 0.03 |
| MVMR IVW | 0.02 | 0.02 | 0.02 | 0.09 | 0.09 | 0.09 | 0.03 | 0.03 | 0.03 |
|  | **Coverage (%)** |  |  |  |  |  |  |  |  |
| LMM-MEC | 89 | 96 | 86 | 84 | 89 | 86 | 95 | 88 | 75 |
| MVMR-Egger | 90 | 96 | 87 | 84 | 89 | 86 | 95 | 87 | 74 |
| MVMR IVW | 90 | 96 | 88 | 45 | 46 | 42 | 28 | 20 | 17 |
|  | **Power (%)** |  |  |  |  |  |  |  |  |
| LMM-MEC | 97 | — | 97 | 81 | — | 82 | 94 | — | 58 |
| MVMR-Egger | 97 | — | 96 | 82 | — | 83 | 94 | — | 59 |
| MVMR IVW | 98 | — | 98 | 72 | — | 73 | 100 | — | 22 |

We applied our method to univariate MR methods, as a special case of MVMR. Our method showed comparable performance under no pleiotropy and balanced pleiotropy, and higher coverage rates than existing methods under directional pleiotropy (**Tables S2, S3**). By increasing the sample size from 5000 to 20000, the coverage rates increased to 95% when using independent IVs, while the coverage rates improved to nearly 95% only under conditions of no pleiotropy when using correlated IVs (**Table S4**).

## 4. APPLICATION STUDY TO LIPOPROTEIN CHOLESTEROL BIOMARKERS AND LONGEVITY

We applied our model to examine the causal associations of blood lipoprotein cholesterol biomarkers with longevity for two reasons. First, state-of-the-art technologies have enabled the profiling of correlated lipidomic markers, and these emerging data have served as powerful tools for understanding complex biological systems.^29,30^ We utilized blood lipoprotein data to show the potential wide application of our model to correlated biomarker data. Second, genome-wide summary statistics with lipoprotein cholesterol biomarkers and longevity are available as open resources, which provides an excellent opportunity to investigate this question.

### 4.1. Methods of the Application Study

Summary statistics related to longevity were obtained from a longevity genomics consortium, where longevity was defined as an age >90th survival percentile.^31^ The summary statistics were generated from 11,262 cases who survived at or beyond this age threshold, and 25,483 controls whose age at death or at last contact was at or below the age corresponding to the 60th survival percentile. Summary statistics for lipoprotein cholesterol biomarkers were sourced from the MAGNETIC consortium, generated from GWAS involving 24,925 participants.^32^

The lipoprotein cholesterol biomarkers were measured using a nuclear magnetic resonance (NMR) metabolomics platform (Nightingale Health Ltd., Helsinki, Finland). Ten cholesterol biomarkers were considered, namely, cholesterol in very large high-density lipoprotein (HDL) (XL.HDL.C), cholesterol in large HDL (L.HDL.C), cholesterol in medium HDL (M.HDL.C), cholesterol in large very low-density lipoprotein (VLDL) (L.VLDL.C), cholesterol in medium VLDL (M.VLDL.C), cholesterol in small VLDL (S.VLDL.C), cholesterol in large low-density lipoprotein (LDL) (L.LDL.C), cholesterol in medium LDL (M.LDL.C), cholesterol in small LDL (S.LDL.C), cholesterol in intermediate-density lipoprotein (IDL) (IDL.C).

To evaluate the performance of our model, we selected genetic variants that were strongly associated with at least one of the lipoprotein cholesterol biomarkers from the GWAS (<5×10^-8^, 5×10^-5^).^33^ The F-statistic of an IV was estimated as the square of the coefficient divided by the square of the standard error of the summary statistics. The Conditional F-statistic was calculated using the ‘strength_mvmr’ command.^23^ These genetic variants were pruned to low LD (*ρ*^2^≤0.1) using the SNPclip tool in LDlink.^34^ The correlation between genetic variants, *ρ*, was estimated in two steps. First, *ρ*^2^ was estimated using the LDmatrix Tool in LDlink.^34^ Second, the effective allele frequency (EAF) was used to determine the sign of the square root of LD, with a positive value assigned if the absolute difference in EAF between two genetic variants is smaller than the absolute difference between the EAF of one genetic variant and the minor allele frequency (MAF) of the other genetic variant.

We applied the LMM-MEC to examine the causal associations between cholesterol biomarkers and longevity, where the correlation between summary statistics with lipoprotein cholesterol biomarkers was obtained by calculating pairwise Pearson correlations. In step II, we simulated the validation study for measurement error correction for 100 times. We compared the findings from our approach to those obtained using MVMR-IVW, MVMR-Egger, MVMR-median, and MVMR-PRESSO methods. For estimated causal associations, we corrected for multiple testing using Bonferroni correction and defined significance as a P value < 0.05/10, where 10 was the number of cholesterol biomarkers.

### 4.2. Results of the Application Study

Using genetic variants selected with stringent P values (P < 5×10^-8^), only a few biomarkers were identified as significantly associated with longevity (**Table 3**). By using genetic variants with strong but less stringent P values (P < 5×10^-5^), our model identified large LDL cholesterol (L.LDL.C) significantly inversely associated with longevity. MVMR-IVW, MVMR-Egger, and MVMR-PRESSO methods further identified mediam HDL cholesterol (M.HDL.C) as significantly positively associated with longevity. The MVMR-median method identified far more significant biomarkers than the other methods.

**Table 3.** Causal associations between lipoprotein cholesterol biomarkers and longevity using multivariable Mendelian randomization.

| Cholesterol biomarkers | Mean F statistic | Conditional F | LMM-MEC | MVMR-Egger | MVMR IVW | MVMR median-based | MR-PRESSO |
| --- | --- | --- | --- | --- | --- | --- | --- |
| Genetic variants with P value<5×10<sup>-8</sup> and low linkage disequilibrium ($\rho^2 \leq 0.1$) (No of IVs = 47) | | | | | | | |
| XL.HDL.C | 16.77 | 4.21 | -0.50 (-2.92, 1.92); 0.68 | -0.90 (-2.61, 0.80); 0.30 | -0.71 (-2.19, 0.78); 0.35 | -0.95 (-1.86, -0.05); 0.04 | -0.68 (-2.09, 0.74); 0.36 |
| L.HDL.C | 27.92 | 4.31 | -0.31 (-2.85, 2.24); 0.81 | 0.90 (-0.92, 2.73); 0.33 | 0.75 (-0.95, 2.46); 0.39 | 0.94 (0.07, 1.82); 0.03 | 0.65 (-0.96, 2.25); 0.43 |
| M.HDL.C | 10.66 | 3.69 | -0.69 (-3.12, 1.74); 0.58 | 0.11 (-0.79, 1.01); 0.81 | 0.17 (-0.69, 1.02); 0.70 | 0.14 (-0.44, 0.72); 0.64 | 0.27 (-0.51, 1.05); 0.51 |
| L.VLDL.C | 10.05 | 2.25 | 1.07 (-1.26, 3.39); 0.37 | 1.73 (-0.43, 3.89); 0.12 | 1.78 (-0.35, 3.91); 0.10 | **1.57 (0.56, 2.59); 0.002** | 1.66 (-0.41, 3.73); 0.13 |
| M.VLDL.C | 13.54 | 10.55 | -1.56 (-4.67, 1.56); 0.33 | -1.78 (-4.24, 0.67); 0.15 | -1.84 (-4.26, 0.57); 0.14 | **-1.66 (-2.75, -0.58); 0.003** | -1.65 (-4.07, 0.76); 0.19 |
| S.VLDL.C | 19.34 | 2.76 | 0.71 (-2.62, 4.03); 0.68 | 0.86 (-1.80, 3.52); 0.53 | 0.83 (-1.80, 3.46); 0.54 | 0.73 (-0.43, 1.90); 0.22 | 0.66 (-1.87, 3.19); 0.61 |
| L.LDL.C | 38.44 | 41.16 | -1.64 (-6.48, 3.21); 0.51 | -1.04 (-5.71, 3.62); 0.66 | -1.10 (-5.71, 3.50); 0.64 | 1.29 (0.02, 2.57); 0.05 | -0.92 (-5.48, 3.65); 0.70 |
| M.LDL.C | 36.35 | 10.64 | 3.96 (-2.11, 10.03); 0.20 | 2.65 (-1.99, 7.28); 0.26 | 2.83 (-1.69, 7.36); 0.22 | -0.17 (-1.35, 1.02); 0.78 | 2.51 (-2.01, 7.02); 0.28 |
| S.LDL.C | 31.67 | 3.88 | -3.58 (-7.79, 0.63); 0.10 | -2.65 (-5.93, 0.63); 0.11 | -2.85 (-5.99, 0.29); 0.08 | -1.13 (-2.30, 0.03); 0.06 | -2.69 (-5.69, 0.31); 0.09 |
| IDL.C | 36.66 | 13.02 | 0.93 (-2.85, 4.70); 0.63 | 0.61 (-1.99, 3.22); 0.64 | 0.68 (-1.88, 3.25); 0.60 | -0.32 (-1.43, 0.79); 0.57 | 0.76 (-1.82, 3.33); 0.57 |
| Genetic variants with P value<5×10<sup>-5</sup> and low linkage disequilibrium ($\rho^2 \leq 0.1$) (No of IVs = 739) | | | | | | | |
| XL.HDL.C | 5.37 | 3.33 | 0.21 (-0.06, 0.49); 0.13 | 0.32 (0.07, 0.56); 0.01 | 0.31 (0.08, 0.54); 0.01 | **0.43 (0.27, 0.58); <0.001** | 0.30 (0.07, 0.53); 0.010 |
| L.HDL.C | 8.03 | 4.09 | -0.36 (-0.68, -0.05); 0.02 | -0.23 (-0.51, 0.06); 0.12 | -0.23 (-0.51, 0.06); 0.12 | **-0.33 (-0.50, -0.16); <0.001** | -0.23 (-0.51, 0.05); 0.107 |
| M.HDL.C | 4.53 | 2.87 | 0.17 (-0.05, 0.39); 0.13 | **0.29 (0.12, 0.46); 0.001** | **0.29 (0.12, 0.46); 0.001** | **0.32 (0.17, 0.46); <0.001** | **0.32 (0.15, 0.49); 0.000** |
| L.VLDL.C | 5.52 | 3.42 | -0.37 (-0.69, -0.05); 0.02 | -0.38 (-0.68, -0.07); 0.02 | -0.38 (-0.68, -0.07); 0.02 | **-0.39 (-0.57, -0.21); <0.001** | -0.31 (-0.61, 0.00); 0.049 |
| M.VLDL.C | 7.39 | 6.07 | 0.33 (-0.10, 0.77); 0.14 | 0.26 (-0.16, 0.68); 0.23 | 0.26 (-0.16, 0.68); 0.22 | **0.54 (0.33, 0.75); <0.001** | 0.14 (-0.27, 0.56); 0.497 |
| S.VLDL.C | 8.53 | 7.40 | 0.44 (0.01, 0.86); 0.05 | 0.38 (-0.02, 0.78); 0.06 | 0.38 (-0.02, 0.78); 0.06 | -0.01 (-0.23, 0.21); 0.94 | 0.43 (0.03, 0.82); 0.035 |
| L.LDL.C | 9.98 | 8.18 | **-1.21 (-1.91, -0.52); <0.001** | **-1.24 (-1.97, -0.52); <0.001** | **-1.24 (-1.96, -0.52); <0.001** | **-0.64 (-0.88, -0.41); <0.001** | -1.01 (-1.72, -0.29); 0.006 |
| M.LDL.C | 9.87 | 6.17 | 0.46 (-0.25, 1.16); 0.20 | 0.38 (-0.28, 1.05); 0.26 | 0.38 (-0.28, 1.05); 0.26 | **0.54 (0.31, 0.77); <0.001** | 0.34 (-0.33, 1.01); 0.318 |
| S.LDL.C | 9.51 | 9.17 | -0.04 (-0.48, 0.40); 0.85 | -0.08 (-0.50, 0.34); 0.70 | -0.08 (-0.50, 0.33); 0.70 | -0.30 (-0.52, -0.08); 0.01 | -0.19 (-0.61, 0.22); 0.363 |
| IDL.C | 9.58 | 8.51 | 0.50 (0.07, 0.93); 0.02 | 0.44 (0.06, 0.83); 0.03 | 0.44 (0.05, 0.83); 0.03 | 0.05 (-0.15, 0.25); 0.62 | 0.36 (-0.02, 0.75); 0.065 |

XL.HDL.C: Cholesterol in very large high-density lipoprotein (HDL); L.HDL.C: Cholesterol in large HDL; M.HDL.C: Cholesterol in medium HDL; L.VLDL.C: Cholesterol in large very low-density lipoprotein (VLDL); M.VLDL.C: Cholesterol in medium VLDL; S.VLDL.C: Cholesterol in small VLDL; L.LDL.C: Cholesterol in large low-density lipoprotein (LDL); M.LDL.C: Cholesterol in medium LDL; S.LDL.C: Cholesterol in small LDL; IDL.C: Cholesterol in intermediate-density lipoprotein (IDL)

## 5. DISCUSSION

We propose a summary-data-based MVMR model that accounts for variance in the summary statistics of both risk factors and the disease outcome. When using independent genetic variants as IVs, our method achieved 95% coverage and was robust to pleiotropy. Particularly, our method showed coverage rate comparable to existing MVMR methods under conditions of no pleiotropy and balanced pleiotropy, and exhibited higher coverage rates under directional pleiotropy. When using correlated genetic variants as IVs, our method is more robust to pleiotropy than existing MVMR methods, although the type 1 error inflated for all methods.

The varied robustness of MVMR methods to the pleiotropy with the outcome could be due to the different model algorithms. MVMR-IVW calculates inverse-variance weighted estimates without an intercept term, making it more sensitive to directional pleiotropy.^17^ MVMR-PRESSO identifies genetic variants with heterogenous effects (potentially due to directional pleiotropy with the outcome) as outliers, and leaves out these variants one at a time to estimate causal effects assuming no directional pleiotropy. MVMR-median also assumes no directional pleiotropy and employs a rank inversion technique to generate narrow 95% confidence interval.^17^ This may explain its low coverage rate in the simulation study and the significantly higher number of lipid biomarkers identified compared to other MVMR methods in the application study. Unlike these methods, MVMR-Egger accommodates directional pleiotropy by fitting an intercept term to the model.^17^ Our method also deals with directional pleiotropy by including an intercept term, however, we apply an algorithm for random-effects model which is different from MVMR-Egger. Moreover, our approach simultaneously accounts for variance in summary statistics of risk factors by applying a multiple measurement error correction method. Compared to existing MVMR methods, our method is more robust to pleiotropy and achieves 95% coverage when using independent genetic variants as IVs.

When using correlated genetic variants as IVs, MVMR-IVW and MVMR-Egger account for correlations between genetic variants using generalized weighted linear regression, where the variant correlations are included in the weighting matrix. Our method accounts for the correlation between genetic variants in two steps. In step I, the correlation between the disease summary statistics is approximated by the Pearson correlation between genetic variance according to Burgess *et al* (2016).^24^ Notably, the iteratively re-weighted least squares algorithm we adopt to model random-effects allows for correlation between genetic variants.^18^ In step II, we simulate a dataset based on the summary statistics of risk factors, which follows a multivariate normal distribution with a covariance structure that accounts for both the correlation between genetic variants and the correlation between summary statistics of the risk factors.

Our method is more robust to pleiotropy than existing MVMR methods using correlated genetic variants as IVs, however, the coverage rates were reduced to below 95% for all methods. Studies have suggested that correlation between genetic variants increases type I error in MR-Egger regression.^35^ We further assessed the performance of our method by increasing the sample size of the simulation to 20000. The coverage rates remained below 95%, while the Monte Carlo standard errors of coverage were less than 0.5%, suggesting systematic under coverage.^36^ The inflated type I error of our method as well as existing methods can be explained as follows. When a risk factor or the disease outcome is determined by multiple correlated genetic variants, the summary statistics of a single genetic variant with the risk factor or disease outcome cannot fully represent the true associations between the genetic variant and the phenotypes. Unlike the variances of summary statistics that can be considered as random errors and be corrected, the systematic errors in the mean values of the summary statistics biases the estimated causal effects and are challenging to eliminate.

Our method accounts for variance in summary statistics with risk factors by applying measurement error correction. In a typical measurement error correction, we correct for the observed association for a risk factor with measurement error. Thus, in the validation study, the true risk factor is regressed on the observed risk factor with measurement error to obtain 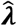. This typically strengthens the association between a risk factor and the outcome for single measurement error correction, as random measurement error in the independent variable attenuates the coefficient 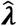 towards null.^37^ However, in step I of our method, we obtain the causal effects for the true 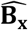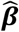, the mean values of 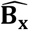, is an unbiased estimation of the true **B**_**x**_), and in step II, our goal is to estimate the corrected causal effects using the observed 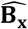 (i.e., 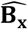 with measurement error). Thus, 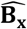 with measurement error (simulated as 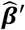) is regressed on 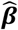 to obtain 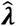. In single measurement error correction, this does not affect effect size but widens the 95% confidence interval, as random measurement error in the dependent variable does not bias the coefficient 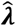 but increases its standard error.^37^ However, in multiple measurement error correction, the value of 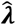 becomes unpredictable due to the correlated measurement error between genetic variants and between risk factors.

In previous MR studies, a stringent threshold of p < 5×10^-8^ was used to select very strong IVs, which may be due to two reasons. First, this threshold helps minimize pleiotropy with the outcome. Second, it minimizes the bias caused by ignoring the uncertainty in summary statistics with risk factor under the NOME assumption. However, this stringent threshold may reduce the power of MVMR, particularly when only a limited number of genetic variants are selected, and the summary statistics of risk factors are correlated. In the application study, we relaxed the threshold to strong but less stringent cutoff (p < 5×10^-5^), which allowed for the inclusion of more genetic variants. MVMR-Egger and MVMR IVW identified medium HDL-c positively associated with longevity and large LDL-c inversely associated. However, our method only identified large LDL-c inversely associated with the likelihood of achieving longevity. The likely reason is that the measurement error correction in our method widened the 95% CI, leading to more conservative findings.

From a biological perspective, a high level of large LDL cholesterol indicates slowed clearance of cholesterol by the liver and is considered a risk factor of cardiovascular disease and reduced longevity.^38^ However, our results need to be interpreted with caution, as using variants that are less strongly (still considered as strong IV) associated with risk factors can create two potential issues. First, it may reduce the values of F-statistics and exacerbate weak instrument bias. Second, it may increase the bias due to pleiotropy, as genetic variants that are weakly associated with an exposure of interest are often thought to be more likely to be pleiotropic. Although our method relaxes the NOME assumption, it still needs strong IV assumption, an inherent feature of instrumental variable analysis.^39,40^ While our method is robust to pleiotropy, the InSIDE assumption is still required, which is very difficult to guarantee in practice that this assumption holds (for example, the genetic variants have effects on an unknown confounder U).

In summary, we have developed a summary-data-based MVMR that accounts for the variance not only in the summary statistics of risk factors, but also in the outcome. Our method is robust to pleiotropy with the disease outcome and can be used to examine the causal associations of multiple exposures with the disease outcome.

## Data Availability

All data produced in the present study are available upon reasonable request to the authors

## ACKNOWLEDGMENTS

This work was supported by grant R21 HG012365 from the National Institutes of Health. The authors would like to acknowledge the highly constructive comments on the manuscript provided by Dr. Daniel I. Chasman (Division of Preventive Medicine, Department of Medicine, Brigham and Women’s Hospital and Harvard Medical School, Boston, MA, USA).

## DATA AVAILABILITY STATEMENT

‘LMM_MEC’ R macro and the code used for the simulations and the application are available at the Github repository (https://github.com/mingding-hsph/LMM_MEC). Summary statistics with longevity is openly available from Longevity Genomics website (https://www.longevitygenomics.org/downloads), and summary statistics with lipoprotein cholesterol biomarkers is also openly available from GWAS Catalog (https://www.ebi.ac.uk/gwas/publications/27005778).

---

**Table S1.** A simulation study estimating causal associations between positive (X_1_), null (X_2_), and negative (X_3_) risk factors and outcome (Y) using LMM-MEC with different sample sizes.

| Sample size | No pleiotropy with outcome |  |  | Balanced pleiotropy, InSIDE satisfied |  |  | Directional pleiotropy, InSIDE satisfied |  |  |
| --- | --- | --- | --- | --- | --- | --- | --- | --- | --- |
| | $k_1=0.1$ | $k_2=0$ | $k_3=-0.1$ | $k_1=0.3$ | $k_2=0$ | $k_3=-0.3$ | $k_1=0.1$ | $k_2=0$ | $k_3=-0.1$ |
| No LD ($\rho^2 = 0$) | | | | | | | | | |
|  | Estimated mean value |  |  |  |  |  |  |  |  |
| 2000 | 0.09 | 0.00 | -0.09 | 0.26 | 0.00 | -0.27 | 0.09 | 0.00 | -0.09 |
| 10000 | 0.10 | 0.00 | -0.10 | 0.29 | 0.00 | -0.30 | 0.10 | 0.00 | -0.10 |
| 20000 | 0.10 | 0.00 | -0.10 | 0.29 | 0.00 | -0.30 | 0.10 | 0.00 | -0.10 |
|  | Estimated standard error |  |  |  |  |  |  |  |  |
| 2000 | 0.02 | 0.02 | 0.02 | 0.08 | 0.08 | 0.08 | 0.02 | 0.02 | 0.02 |
| 10000 | 0.01 | 0.01 | 0.01 | 0.07 | 0.07 | 0.07 | 0.01 | 0.01 | 0.01 |
| 20000 | 0.01 | 0.01 | 0.01 | 0.07 | 0.07 | 0.07 | 0.01 | 0.01 | 0.01 |
|  | Coverage (%) |  |  |  |  |  |  |  |  |
| 2000 | 91 | 97 | 90 | 94 | 95 | 94 | 92 | 96 | 93 |
| 10000 | 94 | 96 | 93 | 94 | 95 | 95 | 92 | 96 | 95 |
| 20000 | 94 | 95 | 94 | 94 | 94 | 95 | 95 | 96 | 95 |
|  | Monte Carlo standard errors of coverage (%) * |  |  |  |  |  |  |  |  |
| 2000 | 0.64 | 0.38 | 0.67 | 0.53 | 0.49 | 0.53 | 0.61 | 0.44 | 0.57 |
| 10000 | 0.24 | 0.20 | 0.26 | 0.24 | 0.22 | 0.22 | 0.27 | 0.20 | 0.22 |
| 20000 | 0.17 | 0.15 | 0.17 | 0.17 | 0.17 | 0.15 | 0.15 | 0.14 | 0.15 |
|  | Power (%) |  |  |  |  |  |  |  |  |
| 2000 | 9 | — | 100 | 94 | — | 94 | 99 | — | 99 |
| 10000 | 100 | — | 100 | 98 | — | 98 | 100 | — | 100 |
| 20000 | 100 | — | 100 | 98 | — | 99 | 100 | — | 100 |
| Low LD ($0 < \rho^2 \leq 0.1$) | | | | | | | | | |
|  | Estimated mean value |  |  |  |  |  |  |  |  |
| 2000 | 0.09 | 0.00 | -0.09 | 0.26 | 0.00 | -0.26 | 0.10 | 0.01 | -0.08 |
| 10000 | 0.10 | 0.00 | -0.10 | 0.29 | 0.00 | -0.29 | 0.11 | 0.01 | -0.08 |
| 20000 | 0.10 | 0.00 | -0.10 | 0.29 | 0.00 | -0.30 | 0.11 | 0.01 | -0.08 |
|  | Estimated standard error |  |  |  |  |  |  |  |  |
| 2000 | 0.02 | 0.02 | 0.02 | 0.08 | 0.08 | 0.08 | 0.02 | 0.02 | 0.02 |
| 10000 | 0.01 | 0.01 | 0.01 | 0.07 | 0.07 | 0.07 | 0.01 | 0.01 | 0.01 |
| 20000 | 0.01 | 0.01 | 0.01 | 0.07 | 0.07 | 0.07 | 0.01 | 0.01 | 0.01 |
|  | Coverage (%) |  |  |  |  |  |  |  |  |
| 2000 | 91 | 95 | 91 | 91 | 92 | 91 | 95 | 93 | 82 |
| 10000 | 95 | 95 | 94 | 90 | 91 | 90 | 87 | 83 | 76 |
| 20000 | 94 | 95 | 93 | 90 | 92 | 90 | 79 | 76 | 71 |
|  | Monte Carlo standard errors of coverage (%) * |  |  |  |  |  |  |  |  |
| 2000 | 0.64 | 0.49 | 0.64 | 0.64 | 0.61 | 0.64 | 0.49 | 0.57 | 0.86 |
| 10000 | 0.22 | 0.22 | 0.24 | 0.30 | 0.29 | 0.30 | 0.34 | 0.38 | 0.43 |
| 20000 | 0.17 | 0.15 | 0.18 | 0.21 | 0.19 | 0.21 | 0.29 | 0.30 | 0.32 |
|  | Power (%) |  |  |  |  |  |  |  |  |
| 2000 | 99 | — | 99 | 89 | — | 89 | 98 | — | 90 |
| 10000 | 100 | — | 100 | 95 | — | 94 | 100 | — | 100 |
| 20000 | 100 | — | 100 | 95 | — | 97 | 100 | — | 100 |
| Moderate LD ($0.1 < \rho^2 \leq 0.3$) | | | | | | | | | |
|  | **Estimated mean value** |  |  |  |  |  |  |  |  |
| 2000 | 0.09 | 0.00 | -0.09 | 0.26 | -0.01 | -0.26 | 0.11 | 0.02 | -0.06 |
| 10000 | 0.10 | 0.00 | -0.10 | 0.29 | 0.00 | -0.30 | 0.13 | 0.04 | -0.06 |
| 20000 | 0.10 | 0.00 | -0.10 | 0.31 | 0.00 | -0.29 | 0.13 | 0.04 | -0.06 |
|  | **Estimated standard error** |  |  |  |  |  |  |  |  |
| 2000 | 0.02 | 0.02 | 0.02 | 0.09 | 0.09 | 0.09 | 0.03 | 0.03 | 0.03 |
| 10000 | 0.01 | 0.01 | 0.01 | 0.08 | 0.07 | 0.08 | 0.02 | 0.02 | 0.02 |
| 20000 | 0.01 | 0.01 | 0.01 | 0.07 | 0.07 | 0.07 | 0.02 | 0.02 | 0.02 |
|  | **Coverage (%)** |  |  |  |  |  |  |  |  |
| 2000 | 88 | 95 | 89 | 83 | 91 | 85 | 95 | 90 | 75 |
| 10000 | 93 | 93 | 92 | 78 | 77 | 77 | 59 | 50 | 47 |
| 20000 | 94 | 94 | 93 | 73 | 77 | 75 | 36 | 37 | 37 |
|  | **Monte Carlo standard errors of coverage (%) \*** |  |  |  |  |  |  |  |  |
| 2000 | 0.73 | 0.49 | 0.70 | 0.84 | 0.64 | 0.80 | 0.49 | 0.67 | 0.97 |
| 10000 | 0.26 | 0.26 | 0.27 | 0.41 | 0.42 | 0.42 | 0.49 | 0.50 | 0.50 |
| 20000 | 0.17 | 0.17 | 0.18 | 0.31 | 0.30 | 0.31 | 0.34 | 0.34 | 0.34 |
|  | **Power (%)** |  |  |  |  |  |  |  |  |
| 2000 | 96 | — | 98 | 80 | — | 80 | 95 | — | 54 |
| 10000 | 100 | — | 100 | 86 | — | 87 | 100 | — | 88 |
| 20000 | 100 | — | 100 | 89 | — | 88 | 100 | — | 94 |

\*Monte Carlo standard errors of coverage is estimated as $\sqrt{\frac{\text{coverage}(1-\text{coverage})}{\text{sample size}}}$ .

**Table S2.** A simulation study estimating causal association between univariate risk factor (X) and outcome (Y) with positive, null, or negative effect on disease, under no linkage disequilibrium (*ρ*^2^ = 0) between genetic variants.

|  | No pleiotropy with outcome |  |  | Balanced pleiotropy, InSIDE satisfied |  |  | Directional pleiotropy, InSIDE satisfied |  |  |
| --- | --- | --- | --- | --- | --- | --- | --- | --- | --- |
| | $k = 0.1$ | $k = 0$ | $k = -0.1$ | $k = 0.3$ | $k = 0$ | $k = -0.3$ | $k = 0.1$ | $k = 0$ | $k = -0.1$ |
|  | **Estimated mean value** |  |  |  |  |  |  |  |  |
| LMM-MEC | 0.09 | 0.00 | -0.09 | 0.26 | 0.00 | -0.27 | 0.09 | 0.02 | -0.03 |
| MR-Egger | 0.06 | 0.00 | -0.06 | 0.18 | 0.00 | -0.17 | 0.06 | 0.02 | -0.01 |
| MR IVW | 0.09 | 0.00 | -0.09 | 0.27 | 0.00 | -0.27 | 0.12 | 0.05 | 0.00 |
| MR-median | 0.08 | 0.00 | -0.08 | 0.26 | 0.00 | -0.26 | 0.12 | 0.05 | 0.00 |
| MR-PRESSO | 0.09 | 0.00 | -0.09 | 0.27 | 0.00 | -0.27 | 0.12 | 0.05 | 0.00 |
| Radial IVW MR | 0.09 | 0.00 | -0.09 | 0.27 | 0.00 | -0.27 | 0.12 | 0.05 | 0.00 |
| Radial Egger MR | 0.06 | 0.00 | -0.06 | 0.19 | 0.00 | -0.18 | 0.07 | 0.02 | -0.01 |
|  | **Estimated standard error** |  |  |  |  |  |  |  |  |
| LMM-MEC | 0.02 | 0.02 | 0.02 | 0.07 | 0.07 | 0.07 | 0.02 | 0.02 | 0.02 |
| MR-Egger | 0.04 | 0.04 | 0.04 | 0.16 | 0.16 | 0.16 | 0.05 | 0.05 | 0.05 |
| MR IVW | 0.02 | 0.02 | 0.02 | 0.07 | 0.07 | 0.07 | 0.02 | 0.02 | 0.02 |
| MR-median | 0.03 | 0.03 | 0.03 | 0.06 | 0.05 | 0.06 | 0.03 | 0.03 | 0.03 |
| MR-PRESSO | 0.02 | 0.02 | 0.02 | 0.07 | 0.07 | 0.07 | 0.02 | 0.02 | 0.02 |
| Radial IVW MR | 0.02 | 0.02 | 0.02 | 0.07 | 0.07 | 0.07 | 0.02 | 0.02 | 0.02 |
| Radial Egger MR | 0.04 | 0.04 | 0.04 | 0.16 | 0.16 | 0.16 | 0.05 | 0.05 | 0.05 |
|  | **Coverage (%)** |  |  |  |  |  |  |  |  |
| LMM-MEC | 90 | 95 | 88 | 93 | 96 | 93 | 89 | 94 | 87 |
| MR-Egger | 82 | 95 | 82 | 88 | 94 | 85 | 90 | 92 | 81 |
| MR IVW | 90 | 95 | 90 | 93 | 94 | 92 | 81 | 67 | 53 |
| MR-median | 93 | 98 | 94 | 71 | 74 | 72 | 89 | 78 | 62 |
| MR-PRESSO | 89 | 94 | 89 | 93 | 94 | 91 | 78 | 65 | 51 |
| Radial IVW MR | 89 | 94 | 89 | 93 | 94 | 91 | 78 | 65 | 51 |
| Radial Egger MR | 80 | 95 | 81 | 89 | 94 | 86 | 91 | 92 | 82 |
|  | **Power (%)** |  |  |  |  |  |  |  |  |
| LMM-MEC | 100 | — | 100 | 96 | — | 96 | 100 | — | 100 |
| MR-Egger | 27 | — | 27 | 21 | — | 20 | 26 | — | 20 |
| MR IVW | 100 | — | 100 | 97 | — | 96 | 100 | — | 84 |
| MR-median | 91 | — | 93 | 94 | — | 94 | 99 | — | 59 |
| MR-PRESSO | 100 | — | 100 | 97 | — | 96 | 100 | — | 81 |
| Radial IVW MR | 100 | — | 100 | 97 | — | 96 | 100 | — | 82 |
| Radial Egger MR | 29 | — | 31 | 22 | — | 22 | 29 | — | 20 |

**Table S3.** A simulation study estimating causal association between univariate risk factor (X) and outcome (Y) with positive, null, or negative effect on disease, under low to moderate linkage disequilibrium (0 < *ρ*^2^ ≤ 0.3) between genetic variants.

|  | No pleiotropy with outcome |  |  | Balanced pleiotropy, InSIDE satisfied |  |  | Directional pleiotropy, InSIDE satisfied |  |  |
| --- | --- | --- | --- | --- | --- | --- | --- | --- | --- |
| | $k = 0.1$ | $k = 0$ | $k = -0.1$ | $k = 0.3$ | $k = 0$ | $k = -0.3$ | $k = 0.1$ | $k = 0$ | $k = -0.1$ |
| Low LD ($0 < \rho^2 \leq 0.1$) | | | | | | | | | |
|  | **Estimated mean value** |  |  |  |  |  |  |  |  |
| LMM-MEC | 0.08 | 0.00 | -0.08 | 0.25 | 0.00 | -0.24 | 0.09 | 0.01 | -0.07 |
| MR-Egger | 0.08 | 0.00 | -0.08 | 0.25 | 0.00 | -0.25 | 0.14 | 0.05 | -0.04 |
| MR IVW | 0.10 | 0.00 | -0.10 | 0.31 | 0.00 | -0.29 | 0.42 | 0.32 | 0.22 |
|  | **Estimated standard error** |  |  |  |  |  |  |  |  |
| LMM-MEC | 0.02 | 0.02 | 0.02 | 0.08 | 0.07 | 0.08 | 0.03 | 0.02 | 0.02 |
| MR-Egger | 0.02 | 0.02 | 0.02 | 0.09 | 0.09 | 0.09 | 0.03 | 0.03 | 0.03 |
| MR IVW | 0.01 | 0.01 | 0.01 | 0.05 | 0.04 | 0.04 | 0.02 | 0.02 | 0.02 |
|  | **Coverage (%)** |  |  |  |  |  |  |  |  |
| LMM-MEC | 81 | 95 | 81 | 87 | 92 | 83 | 94 | 92 | 69 |
| MR-Egger | 83 | 92 | 83 | 77 | 79 | 74 | 74 | 70 | 52 |
| MR IVW | 64 | 68 | 67 | 10 | 11 | 12 | 0 | 0 | 0 |
|  | **Power (%)** |  |  |  |  |  |  |  |  |
| LMM-MEC | 98 | — | 98 | 90 | — | 87 | 97 | — | 89 |
| MR-Egger | 97 | — | 98 | 83 | — | 78 | 97 | — | 75 |
| MR IVW | 100 | — | 100 | 91 | — | 90 | 100 | — | 100 |
| Moderate LD ($0.1 < \rho^2 \leq 0.3$) | | | | | | | | | |
|  | **Estimated mean value** |  |  |  |  |  |  |  |  |
| LMM-MEC | 0.07 | 0.00 | -0.07 | 0.22 | 0.00 | -0.22 | 0.11 | 0.04 | -0.04 |
| MR-Egger | 0.08 | 0.00 | -0.08 | 0.23 | 0.00 | -0.22 | 0.11 | 0.04 | -0.04 |
| MR IVW | 0.10 | 0.00 | -0.10 | 0.31 | -0.02 | -0.31 | 0.49 | 0.39 | 0.29 |
|  | **Estimated standard error** |  |  |  |  |  |  |  |  |
| LMM-MEC | 0.02 | 0.02 | 0.02 | 0.08 | 0.08 | 0.08 | 0.03 | 0.03 | 0.03 |
| MR-Egger | 0.02 | 0.02 | 0.02 | 0.08 | 0.08 | 0.08 | 0.03 | 0.03 | 0.03 |
| MR IVW | 0.01 | 0.01 | 0.01 | 0.04 | 0.04 | 0.04 | 0.02 | 0.02 | 0.02 |
|  | **Coverage (%)** |  |  |  |  |  |  |  |  |
| LMM-MEC | 72 | 95 | 70 | 67 | 81 | 66 | 95 | 74 | 31 |
| MR-Egger | 79 | 97 | 76 | 69 | 79 | 68 | 94 | 73 | 33 |
| MR IVW | 66 | 70 | 71 | 3 | 4 | 3 | 0 | 0 | 0 |
|  | **Power (%)** |  |  |  |  |  |  |  |  |
| LMM-MEC | 94 | — | 95 | 74 | — | 73 | 93 | — | 36 |
| MR-Egger | 94 | — | 94 | 74 | — | 74 | 94 | — | 39 |
| MR IVW | 100 | — | 100 | 97 | — | 97 | 100 | — | 100 |

**Table S4.** A simulation study estimating causal associations between univariate risk factor (X) and outcome (Y) with positive, null, or negative effect on disease using LMM-MEC with different sample sizes.

| Sample size | No pleiotropy with outcome |  |  | Balanced pleiotropy, InSIDE satisfied |  |  | Directional pleiotropy, InSIDE satisfied |  |  |
| --- | --- | --- | --- | --- | --- | --- | --- | --- | --- |
| | $k = 0.1$ | $k = 0$ | $k = -0.1$ | $k = 0.3$ | $k = 0$ | $k = -0.3$ | $k = 0.1$ | $k = 0$ | $k = -0.1$ |
| No LD ($\rho^2 = 0$) | | | | | | | | | |
|  | **Estimated mean value** |  |  |  |  |  |  |  |  |
| 2000 | 0.09 | 0.00 | -0.09 | 0.24 | 0.00 | -0.25 | 0.09 | 0.00 | -0.09 |
| 10000 | 0.10 | 0.00 | -0.10 | 0.29 | 0.00 | -0.28 | 0.10 | 0.00 | -0.10 |
| 20000 | 0.10 | 0.00 | -0.10 | 0.29 | 0.00 | -0.29 | 0.10 | 0.00 | -0.10 |
|  | **Estimated standard error** |  |  |  |  |  |  |  |  |
| 2000 | 0.02 | 0.02 | 0.02 | 0.07 | 0.07 | 0.07 | 0.02 | 0.02 | 0.02 |
| 10000 | 0.01 | 0.01 | 0.01 | 0.07 | 0.07 | 0.07 | 0.01 | 0.01 | 0.01 |
| 20000 | 0.01 | 0.01 | 0.01 | 0.07 | 0.07 | 0.07 | 0.01 | 0.01 | 0.01 |
|  | **Coverage (%)** |  |  |  |  |  |  |  |  |
| 2000 | 88 | 96 | 90 | 92 | 96 | 92 | 90 | 95 | 90 |
| 10000 | 94 | 94 | 94 | 94 | 95 | 95 | 93 | 94 | 92 |
| 20000 | 94 | 96 | 94 | 93 | 96 | 96 | 95 | 96 | 94 |
|  | **Monte Carlo standard errors of coverage (%) \*** |  |  |  |  |  |  |  |  |
| 2000 | 0.73 | 0.44 | 0.67 | 0.61 | 0.44 | 0.61 | 0.67 | 0.49 | 0.67 |
| 10000 | 0.24 | 0.24 | 0.24 | 0.24 | 0.22 | 0.22 | 0.26 | 0.24 | 0.27 |
| 20000 | 0.17 | 0.14 | 0.17 | 0.18 | 0.14 | 0.14 | 0.15 | 0.14 | 0.17 |
|  | **Power (%)** |  |  |  |  |  |  |  |  |
| 2000 | 99 | — | 99 | 95 | — | 95 | 99 | — | 100 |
| 10000 | 100 | — | 100 | 99 | — | 99 | 100 | — | 100 |
| 20000 | 100 | — | 100 | 99 | — | 99 | 100 | — | 100 |
| Low LD ($0 < \rho^2 \leq 0.1$) | | | | | | | | | |
|  | **Estimated mean value** |  |  |  |  |  |  |  |  |
| 2000 | 0.08 | 0.00 | -0.08 | 0.24 | 0.00 | -0.24 | 0.09 | 0.01 | -0.07 |
| 10000 | 0.10 | 0.00 | -0.10 | 0.29 | 0.00 | -0.29 | 0.11 | 0.01 | -0.08 |
| 20000 | 0.10 | 0.00 | -0.10 | 0.29 | 0.00 | -0.29 | 0.11 | 0.01 | -0.08 |
|  | **Estimated standard error** |  |  |  |  |  |  |  |  |
| 2000 | 0.02 | 0.02 | 0.02 | 0.08 | 0.07 | 0.08 | 0.02 | 0.02 | 0.02 |
| 10000 | 0.01 | 0.01 | 0.01 | 0.07 | 0.07 | 0.07 | 0.01 | 0.01 | 0.01 |
| 20000 | 0.01 | 0.01 | 0.01 | 0.07 | 0.07 | 0.07 | 0.01 | 0.01 | 0.01 |
|  | **Coverage (%)** |  |  |  |  |  |  |  |  |
| 2000 | 84 | 96 | 81 | 85 | 93 | 85 | 96 | 92 | 69 |
| 10000 | 92 | 95 | 92 | 91 | 91 | 88 | 87 | 78 | 63 |
| 20000 | 93 | 94 | 93 | 89 | 91 | 91 | 77 | 68 | 58 |
|  | **Monte Carlo standard errors of coverage (%) \*** |  |  |  |  |  |  |  |  |
| 2000 | 0.82 | 0.44 | 0.88 | 0.80 | 0.57 | 0.80 | 0.44 | 0.61 | 1.03 |
| 10000 | 0.27 | 0.22 | 0.27 | 0.29 | 0.29 | 0.32 | 0.34 | 0.41 | 0.48 |
| 20000 | 0.18 | 0.17 | 0.18 | 0.22 | 0.20 | 0.20 | 0.30 | 0.33 | 0.35 |
|  | **Power (%)** |  |  |  |  |  |  |  |  |
| 2000 | 98 | — | 99 | 89 | — | 89 | 98 | — | 88 |
| 10000 | 100 | — | 100 | 96 | — | 94 | 100 | — | 100 |
| 20000 | 100 | — | 100 | 96 | — | 95 | 100 | — | 100 |
| Moderate LD ($0.1 < \rho^2 \leq 0.3$) | | | | | | | | | |
|  | **Estimated mean value** |  |  |  |  |  |  |  |  |
| 2000 | 0.07 | 0.00 | -0.07 | 0.21 | -0.01 | -0.22 | 0.11 | 0.03 | -0.04 |
| 10000 | 0.09 | 0.00 | -0.09 | 0.28 | 0.00 | -0.28 | 0.14 | 0.05 | -0.05 |
| 20000 | 0.10 | 0.00 | -0.10 | 0.29 | 0.00 | -0.29 | 0.15 | 0.05 | -0.05 |
|  | **Estimated standard error** |  |  |  |  |  |  |  |  |
| 2000 | 0.02 | 0.02 | 0.02 | 0.08 | 0.08 | 0.08 | 0.03 | 0.03 | 0.03 |
| 10000 | 0.01 | 0.01 | 0.01 | 0.07 | 0.07 | 0.07 | 0.02 | 0.02 | 0.02 |
| 20000 | 0.01 | 0.01 | 0.01 | 0.07 | 0.07 | 0.07 | 0.02 | 0.02 | 0.02 |
|  | **Coverage (%)** |  |  |  |  |  |  |  |  |
| 2000 | 71 | 95 | 70 | 67 | 81 | 66 | 95 | 75 | 33 |
| 10000 | 87 | 95 | 88 | 68 | 69 | 68 | 45 | 31 | 18 |
| 20000 | 91 | 94 | 89 | 67 | 65 | 69 | 28 | 20 | 16 |
|  | **Monte Carlo standard errors of coverage (%) \*** |  |  |  |  |  |  |  |  |
| 2000 | 1.01 | 0.49 | 1.02 | 1.05 | 0.88 | 1.06 | 0.49 | 0.97 | 1.05 |
| 10000 | 0.34 | 0.22 | 0.32 | 0.47 | 0.46 | 0.47 | 0.50 | 0.46 | 0.38 |
| 20000 | 0.20 | 0.17 | 0.22 | 0.33 | 0.34 | 0.33 | 0.32 | 0.28 | 0.26 |
|  | **Power (%)** |  |  |  |  |  |  |  |  |
| 2000 | 94 | — | 94 | 73 | — | 71 | 94 | — | 34 |
| 10000 | 100 | — | 100 | 85 | — | 83 | 100 | — | 71 |
| 20000 | 100 | — | 100 | 85 | — | 87 | 100 | — | 81 |

\*Monte Carlo standard errors of coverage is estimated as $\sqrt{\frac{\text{coverage}(1-\text{coverage})}{\text{sample size}}}$ .

## Notes

### Competing Interest Statement

The authors have declared no competing interest.

### Funding Statement

NHGRI

### Author Declarations

Summary statistics with longevity is openly available from Longevity Genomics website (https://www.longevitygenomics.org/downloads), and summary statistics with lipoprotein cholesterol biomarkers is also openly available from GWAS Catalog (https://www.ebi.ac.uk/gwas/publications/27005778).

### Summary of Updates

We have made a major revision to the simulation section.

## Reference

1. Bowden J, Davey Smith G, Haycock PC, Burgess S. Consistent Estimation in Mendelian Randomization with Some Invalid Instruments Using a Weighted Median Estimator. Genet Epidemiol 2016;40(4):304–14. DOI: 10.1002/gepi.21965.

2. Verbanck M, Chen CY, Neale B, Do R. Detection of widespread horizontal pleiotropy in causal relationships inferred from Mendelian randomization between complex traits and diseases. Nat Genet 2018;50(5):693–698. DOI: 10.1038/s41588-018-0099-7.

3. Zhu Z, Zheng Z, Zhang F, et al. Causal associations between risk factors and common diseases inferred from GWAS summary data. Nat Commun 2018;9(1):224. DOI: 10.1038/s41467-017-02317-2.

4. Zhu Z, Zhang F, Hu H, et al. Integration of summary data from GWAS and eQTL studies predicts complex trait gene targets. Nat Genet 2016;48(5):481–7. DOI: 10.1038/ng.3538.

5. Burgess S, Thompson SG. Interpreting findings from Mendelian randomization using the MR-Egger method. Eur J Epidemiol 2017;32(5):377–389. DOI: 10.1007/s10654-017-0255-x.

6. Qi G, Chatterjee N. Mendelian randomization analysis using mixture models for robust and efficient estimation of causal effects. Nat Commun 2019;10(1):1941. DOI: 10.1038/s41467-019-09432-2.

7. O’Connor LJ, Price AL. Distinguishing genetic correlation from causation across 52 diseases and complex traits. Nat Genet 2018;50(12):1728–1734. DOI: 10.1038/s41588-018-0255-0.

8. Morrison J, Knoblauch N, Marcus JH, Stephens M, He X. Mendelian randomization accounting for correlated and uncorrelated pleiotropic effects using genome-wide summary statistics. Nat Genet 2020. DOI: 10.1038/s41588-020-0631-4.

9. Bowden J, Del Greco MF, Minelli C, et al. Improving the accuracy of two-sample summary-data Mendelian randomization: moving beyond the NOME assumption. Int J Epidemiol 2019;48(3):728–742. DOI: 10.1093/ije/dyy258.

10. Zhao Q, Wang J, Hemani G, Bowden J, Small DS. Statistical inference in two-sample summary-data Mendelian randomization using robust adjusted profile score. arXiv:180109652 2018.

11. Gleason KJ, Yang F, Chen LS. A robust two-sample transcriptome-wide Mendelian randomization method integrating GWAS with multi-tissue eQTL summary statistics. Genet Epidemiol 2021. DOI: 10.1002/gepi.22380.

12. Burgess S, Thompson DJ, Rees JMB, Day FR, Perry JR, Ong KK. Dissecting Causal Pathways Using Mendelian Randomization with Summarized Genetic Data: Application to Age at Menarche and Risk of Breast Cancer. Genetics 2017;207(2):481–487. DOI: 10.1534/genetics.117.300191.

13. Porcu E, Rueger S, Lepik K, et al. Mendelian randomization integrating GWAS and eQTL data reveals genetic determinants of complex and clinical traits. Nat Commun 2019;10(1):3300. DOI: 10.1038/s41467-019-10936-0.

14. Burgess S, Thompson SG. Multivariable Mendelian randomization: the use of pleiotropic genetic variants to estimate causal effects. Am J Epidemiol 2015;181(4):251–60. DOI: 10.1093/aje/kwu283.

15. Sanderson E, Davey Smith G, Windmeijer F, Bowden J. An examination of multivariable Mendelian randomization in the single-sample and two-sample summary data settings. Int J Epidemiol 2019;48(3):713–727. DOI: 10.1093/ije/dyy262.

16. Rees JMB, Wood AM, Burgess S. Extending the MR-Egger method for multivariable Mendelian randomization to correct for both measured and unmeasured pleiotropy. Stat Med 2017;36(29):4705–4718. DOI: 10.1002/sim.7492.

17. Grant AJ, Burgess S. Pleiotropy robust methods for multivariable Mendelian randomization. Stat Med 2021;40(26):5813–5830. DOI: 10.1002/sim.9156.

18. Berkey CS, Hoaglin DC, Mosteller F, Colditz GA. A random-effects regression model for meta-analysis. Stat Med 1995;14(4):395–411. DOI: 10.1002/sim.4780140406.

19. Willett WC. Chapter 12. Correction for the Eects of Measurement Error. Nutritional Epidemiology (3rd edn) 2012:287–304.

20. Rosner B, Spiegelman D, Willett WC. Correction of logistic regression relative risk estimates and confidence intervals for measurement error: the case of multiple covariates measured with error. Am J Epidemiol 1990;132(4):734–45. DOI: 10.1093/oxfordjournals.aje.a115715.

21. Thomas D, Stram D, Dwyer J. Exposure measurement error: influence on exposure-disease. Relationships and methods of correction. Annu Rev Public Health 1993;14:69–93. DOI: 10.1146/annurev.pu.14.050193.000441.

22. Keogh RH, White IR. A toolkit for measurement error correction, with a focus on nutritional epidemiology. Stat Med 2014;33(12):2137–55. DOI: 10.1002/sim.6095.

23. Sanderson E, Spiller W, Bowden J. Testing and correcting for weak and pleiotropic instruments in two-sample multivariable Mendelian randomization. Stat Med 2021;40(25):5434–5452. DOI: 10.1002/sim.9133.

24. Burgess S, Dudbridge F, Thompson SG. Combining information on multiple instrumental variables in Mendelian randomization: comparison of allele score and summarized data methods. Stat Med 2016;35(11):1880–906. DOI: 10.1002/sim.6835.

25. Higgins JP, Thompson SG. Quantifying heterogeneity in a meta-analysis. Stat Med 2002;21(11):1539–58. DOI: 10.1002/sim.1186.

26. Little RJA, Rubin DB. Statistical Analysis with Missing Data, New York: John Wiley & Sons, Inc. 1987.

27. Proof Verification: Joint variance of the product of a random matrix with a random vector. https://stats.stackexchange.com/questions/420032/proof-verification-joint-variance-of-the-product-of-a-random-matrix-with-a-rand. Accessed May, 2021.

28. Broadbent JR, Foley CN, Grant AJ, Mason AM, Staley JR, Burgess S. MendelianRandomization v0.5.0: updates to an R package for performing Mendelian randomization analyses using summarized data. Wellcome Open Res 2020;5:252. DOI: 10.12688/wellcomeopenres.16374.2.

29. Joshi A, Rienks M, Theofilatos K, Mayr M. Systems biology in cardiovascular disease: a multiomics approach. Nat Rev Cardiol 2021;18(5):313–330. DOI: 10.1038/s41569-020-00477-1.

30. Ding M, Zeleznik OA, Guasch-Ferre M, et al. Metabolome-Wide Association Study of the Relationship Between Habitual Physical Activity and Plasma Metabolite Levels. Am J Epidemiol 2019;188(11):1932–1943. DOI: 10.1093/aje/kwz171.

31. Deelen J, Evans DS, Arking DE, et al. A meta-analysis of genome-wide association studies identifies multiple longevity genes. Nat Commun 2019;10(1):3669. DOI: 10.1038/s41467-019-11558-2.

32. Kettunen J, Demirkan A, Wurtz P, et al. Genome-wide study for circulating metabolites identifies 62 loci and reveals novel systemic effects of LPA. Nat Commun 2016;7:11122. DOI: 10.1038/ncomms11122.

33. Burgess S, Butterworth A, Thompson SG. Mendelian randomization analysis with multiple genetic variants using summarized data. Genet Epidemiol 2013;37(7):658–65. DOI: 10.1002/gepi.21758.

34. Machiela MJ, Chanock SJ. LDlink: a web-based application for exploring population-specific haplotype structure and linking correlated alleles of possible functional variants. Bioinformatics 2015;31(21):3555–7. DOI: 10.1093/bioinformatics/btv402.

35. Barfield R, Feng H, Gusev A, et al. Transcriptome-wide association studies accounting for colocalization using Egger regression. Genet Epidemiol 2018;42(5):418–433. DOI: 10.1002/gepi.22131.

36. Morris TP, White IR, Crowther MJ. Using simulation studies to evaluate statistical methods. Stat Med 2019;38(11):2074–2102. DOI: 10.1002/sim.8086.

37. Hutcheon JA, Chiolero A, Hanley JA. Random measurement error and regression dilution bias. BMJ 2010;340:c2289. DOI: 10.1136/bmj.c2289.

38. Wadhera RK, Steen DL, Khan I, Giugliano RP, Foody JM. A review of low-density lipoprotein cholesterol, treatment strategies, and its impact on cardiovascular disease morbidity and mortality. J Clin Lipidol 2016;10(3):472–89. DOI: 10.1016/j.jacl.2015.11.010.

39. Angrist JD, Imbens GW. Two-stage least squares estimation of average causal effects in models with variable treatment intensity. JASA 1995;90:431– 442.

40. Angrist I, Rubin DB. Identification of causal effects using instrumental variables. JASA 1996;91:444–455.

